# BioDeformUNet: A Deep Learning Model for Biomechanically Informed Liver Image Registration

**DOI:** 10.64898/2026.08.03.26359612

**Authors:** Xinyue Zhang, Caleb S. O’Connor, Austin Castelo, McKell E. Woodland, Bilel Daoud, Iwan Paolucci, Jessica Albuquerque, Mais Altaie, Noreen Siddiqi, Ankit B. Patel, Bruno C. Odisio, Kristy K. Brock

## Abstract

**Purpose:** To build a 3D U-Net model, BioDeformUNet, to predict the liver’s deformation vector field (DVF) in near real-time, for efficient intra-procedural evaluation of the minimal ablative margin (MAM).

**Materials and Methods:** This retrospective study included 170 contrast-enhanced computed tomography (CECT) image pairs from 157 patients who underwent liver ablation treatment between 2020-2024. Each data instance included one pre-ablation CECT (pre-CECT) and one post-ablation CECT (post-CECT). BioDeformUNet was trained under the guidance of DVFs generated by a biomechanical model-based deformable image registration (DIR) algorithm using a loss function that focused on large liver deformations. Data were split patient-wise into training (92-93 patients), validation (23-24 patients), and testing sets (42 patients). We compared our method’s performance with two deep learning-based DIR methods: VoxelMorph and VFA. Evaluation metrics included: target registration error (TRE), Dice similarity coefficient (DSC), Minimum Ablation Margin (MAM), and inference time. For BioDeformUNet, we additionally evaluated the accuracy of the deformed tumor center-of-mass mapping by comparing the predicted tumor center location with that generated by Morfeus. A mapping error less than 3.0 mm (corresponding to the voxel size) was considered accurate. We used the Wilcoxon signed-rank test to assess the significancy of each test result. Our code is available at https://github.com/XinyueZhang831/BioDeformUNET.

**Results:** The TRE of BioDeformUNet was not significantly different from Morfeus (3.31±1.17 BioDeformUNet; 3.23±1.06 Morfeus; p-value=0.41). The BioDeformUNet DVF magnitude was within 3.0 mm of Morfeus DVF for an average of 91.9 ± 17.6% (mean ± SD) of the voxels. Tumor mapping errors greater than 3.0 mm occurred in only 8 cases. The inference time of BioDeformUNet was 0.6s per image pair, 0.2s for VoxelMorph, 0.3s for VFA, and 20.2s for Morfeus.

**Conclusion:** BioDeformUNet achieved a similar performance to the biomechanical model-based algorithm but required fewer computational operations, resulting in a 34× speedup in DVF computation.

## Introduction

Liver cancer is the sixth most common cancer and the third leading cause of cancer death worldwide^1^. Image-guided tumor ablation is a treatment option for select patients ^2–5^. In this technique, a probe is inserted into the target tumor to deliver energy to destroy it^6^.

However, accurately positioning the ablation probe within the tumor and ensuring an adequate ablation margin is challenging due to the lack of visualization of the tumor on non-contrast-enhanced computed tomography (CT) images, the lack of conspicuity of the tumor on post-ablation contrast-enhanced CT (CECT), and deformation of the liver over the course of treatment^7^. To address this issue, image registration has been used to map the tumor from the CECT at the start of treatment to the ablation probe–positioning image CECT and post-ablation image ^8^. Clinicians rely on visual mapping, rigid registration, or intensity-based deformable registration (DIR) to map the tumor from pre-CECT to post-CECT. A biomechanical model-based DIR algorithm, Morfeus, has been shown in single and multi-institutional retrospective studies and a randomized phase II study to significantly improve the quantification of the minimum ablation margin (MAM) compared to rigid registration and visual assessment, respectively^2,9^.

While biomechanical DIR demonstrates improved performance compared to intensity-based image registration for liver ablation assessment ^7,10,11^, it requires the full liver to be within the image field of view (FOV) and is a multi-step process that poses constraints on its use^11^. One image registration task requires an average of 3-5 minutes, including AI-based segmentation of the liver and tumor (or ablation zone), generation of the finite element model (FEM), and finite element analysis (FEA). Improving this efficiency will enable rapid DIR for near-real-time applications and improve flexibility in the workflow.

Deep learning may improve algorithm efficiency compared to biomechanical DIR^12^, replacing the FEM and FEA components. Several publicly available unsupervised deep learning-based models for medical images achieve high accuracy in aligning the overall shape of structures. VoxelMorph^13^, for example, is one of the most widely used deep learning models for organ shape deformation, achieving high global accuracy by leveraging voxel intensity similarity. More recent approaches, especially transformer and transformer-inspired models (e.g. TransMorph^14^, VFA^15^), have further improved registration performance. However, because most methods are driven by image similarity, which does not enforce biomechanical consistency, they may not guarantee accurate alignment of internal anatomical features. This limitation is important for some downstream clinical tasks, such as the MAM assessment, an indicator of successful tumor ablation treatment. This study addresses this gap by training a supervised deep learning model to learn biomechanical deformation behaviors and evaluating its consistency with a biomechanical reference. VoxelMorph and VFA were selected as the representative baselines for similarity-driven and attention-based (transformer-inspired) registration methods.

In this retrospective study, we hypothesized that a deep learning algorithm, trained on the results of a biomechanical model-based algorithm instead of image similarity metrics, can perform with an accuracy not significantly different from the biomechanical-based DIR algorithm^11^ with reduced run time. BioDeformUNet is trained to learn the registration behavioral patterns inherent in the DVF generated by Morfeus ^11^.

## Materials and Methods

### Data Use Declaration

Anonymized images were retrospectively obtained from The University of Texas MD Anderson Cancer Center under IRB-approved protocols (PA19-0213, 2021-0340, 2023-0083 and 2024-0340).

### Dataset

We conducted our retrospective study using CT image data from The University of Texas MD Anderson Cancer Center, comprising 170 image pairs from 157 patients (98 men and 59 women, aged 21 to 83 years) who underwent liver ablation from COVER-ALL trial, a clinical cohort treated using CAS-One and Epione systems, and a prospective study for AI model validation from 2020 June to 2024 December. In our dataset, 12 patients had multiple image pairs. The dataset included contrast enhanced CT (CECT) images acquired before the ablation procedure to locate the tumor (pre-CECT), CECT images acquired after the intraprocedural phase to confirm if MAM was achieved (post-CECT), liver masks for pre-CECTs and post-CECTs, and DVFs between the two. CECTs were acquired using standard liver imaging protocols, including contrast-enhanced portal venous phase imaging with a slice thickness of 3 mm. The DVFs were generated using the biomechanical DIR algorithm Morfeus implemented and validated in a commercial radiotherapy treatment planning system (RayStation 11B, RaySearch Laboratories, Stockholm, Sweden^16^) and were used as the ground truth. The DVF generation was performed after probe removal; therefore, it represents the deformation between the pre-ablation and post-ablation states. We use patient level 5-fold cross-validation for model training and selection, with a separate, randomly-selected fixed testing set of 43 data instances for evaluation, where no patient appears in more than one set. The details of the data are shown in Figure 1. In a previous study, an AI model extracted the pre-CECT, post-CECT liver masks, ablation zone, and tumor segmentation ^17^. These outputs were reviewed and, if needed, edited by three trained radiologists with 7 years of experience as part of the clinical process. Tumor localization and MAM assessment were also confirmed by these radiologists.

**Figure 1.**
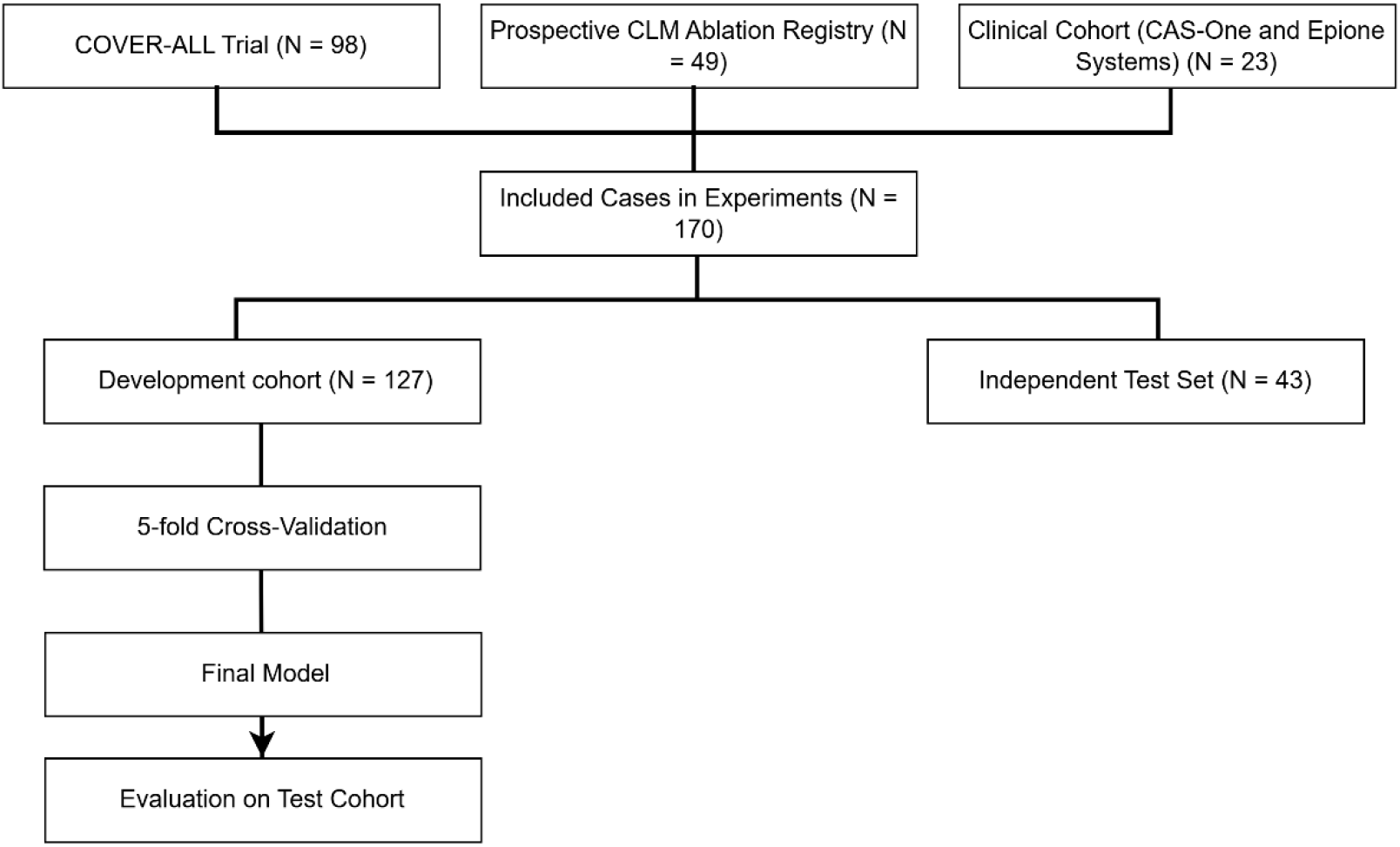
Overview of data sources.

The data preprocessing procedures included the following steps: the CT scans were resampled to have the same voxel spacing (1.14, 1.14, 2.5) and image origin across all datasets; rigid registration was applied on the basis of the geometric center of the liver contours to correct for global shifts between the pre-CECT and post-CECT; CT scans were normalized on the basis of 50 to 350 Hounsfield units^18^, and linearly scaled to [0, 1]; a “liver box” which contains both the pre-CECT liver and post-CECT liver anatomy while preserving their relative positions; and, finally, voxels outside of the liver region were excluded from further analysis. We used the package SimpleITK ^19–21^ to perform the delineated data preprocessing steps. Details of the rigid registration are provided in Supplement 1.

Data augmentation was used to expand the number of training cases and increase the range of initial spatial misalignment so the model could be exposed to cases with substantial displacement and improve performance in large-deformation regions. Additional post-CECT, post-liver mask, and DVF scans were generated by modifying the original DVF. If the maximal displacement in any of the x-, y-, or z-directions was between 5 and 10 mm, the post-CECT and post-liver masks would be shifted along that direction by the difference between 10 mm and the observed deformation. This resulted in more cases with a maximum deformation of 10 mm. No augmentation was performed in the test set. Because the augmentation was applied to the training set across different folds, the number of augmented instances varied across folds.

Due to VRAM constraints, all image data were cropped to the liver box, and zero padding was applied to dimensions 128 (slice direction) x 256 x 256 (in-plane).

### Model

BioDeformUNet, a supervised 3D U-Net model, was constructed with TensorFlow to predict the DVF between pre-CECT and post-CECT scans. The model structure is shown in Fig 2. We used 1e-5 as the learning rate and Adam as the optimizer, selected based on empirical observations and their widespread use in similar deep learning-based medical image registration studies^13,22,23^. The model had four levels and 16 initial filters, two 3D convolutions at each level of the downsampler, one 3D convolution at the upsampler, and a skip connection between the downsampler and upsampler at each level. After each convolution, a batch normalization layer was applied, followed by a Leaky ReLU activation function. We used max pooling to downsample at each level. The training procedure was designed to run for 1500 epochs to ensure sufficient training and to observe convergent behavior over time. Early stopping was used to terminate training if no improvement was observed after 500 epochs; the validation loss determined the best epoch. Our code is available at https://github.com/XinyueZhang831/BioDeformUNET.

**Figure 2.**
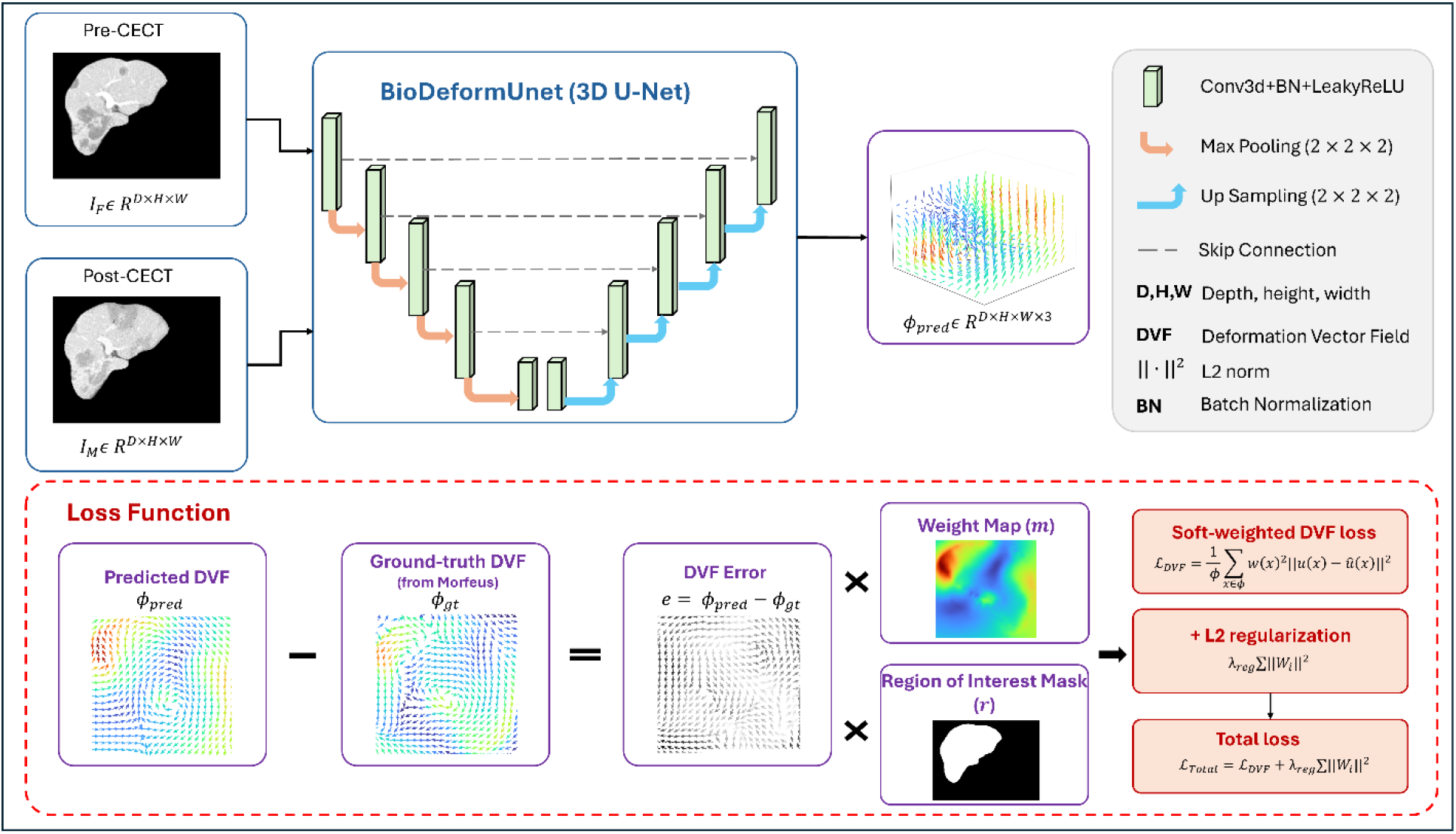
Model structure.

Our training and evaluation processes were executed on four NVIDIA A100 GPUs with 40 GB.

### Loss Function

In our study, we built a task-specific MSE loss function that uses a regional deformation-driven weight (*w*) to improve performance in regions that undergo large deformation. A supervised 3D U-Net model was trained using the ground-truth DVF generated by Morfeus. Let *x* ∈ *φ* denote the voxel location in the DVF, where *φ* represents the number of voxels involved in the loss function, *u*(*x*) is the ground truth deformation and *û(x*) is the predicted deformation at the voxel x.

The weighting term *w*(*x*) was constructed in two parts. First, a region-of-interest (ROI) mask *r*(*x*), obtained from the pre-CT, was applied to focus the loss computation on the liver region, so MSE computes errors only within the liver. Second, a normalized DVF magnitude map *m*(*x*) is used to assign higher weights to regions experiencing large deformation. The resulting DVF loss is defined as

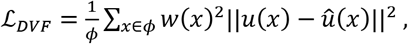

where

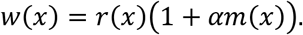

*α* is a fixed hyperparameter controlling the strength of *w*(*x*), and *m*(*x*) is the normalized magnitude of the ground truth DVF, defined as:

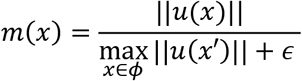

with *ε* added for numerical stability.

In addition, an L2 regularization term is used to improve generalization and prevent overfitting. The overall loss function is therefore

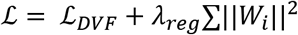

where *W_i_* is the convolution kernel weights of the layer *i*, and *λ_reg_* is the regularization coefficient used to prevent overfitting.

After a hyperparameter search, *α* was set to 0.25, and *λ_reg_* was set to 0.05.

Details of the loss function selection are in Supplement 2.

### Evaluation

We first evaluated the agreement between the deep learning methods and the Morfeus reference standard using the following metrics:

I Tumor localization test: compare the Euclidean distance between the Morfeus reference and the predicted deformed tumor centroid. More detailed information is available in Supplement 3.
II Target Registration Error (TRE): use anatomical landmarks that were automatically identified using a previously validated algorithm,^24^ to calculate the TRE. TRE is the Euclidean distance between corresponding vascular bifurcation landmarks. Figure 3 demonstrates an example of the landmark appearance and shows how the deformable registration algorithm transforms the landmark to align with the target location. Test cases were included in the TRE analysis if they had at least 3 bifurcation points extracted by the automated algorithm, yielding 35 cases.
III For BioDeformUNet, voxel-wise displacement accuracy was further compared with the reference standard. This metric was only reported for BioDeformUNet because it was trained to approximate the Morfeus DVF. Other methods were optimized for image similarity and were not directly comparable. This metric shows the percentage of voxels with an error below the 3-mm threshold. The threshold was set based on a voxel diameter of (1.14, 1.14, 2.5).

Biomechanical DIR methods have been shown to reliably map tumor location^9^, supporting the use of Morfeus as a reference standard in this study.

We further compared the performance of BioDeformUNet to that of VoxelMorph^13^ and VFA^15^, each trained with the same dataset as BioD. Our training approach was based on the fundamental code and parameters included in the GitHub tutorials^12,14^. We further evaluated cross-method performance using the following metrics:

IV Tumor Volume Change: We compared the tumor volume change using the size difference between the tumor size on the pre-CECT and the tumor size after deformation with predicted deformations, defined as the absolute difference and normalized by the tumor size from the pre-CECT. The tumor volume should not change when the anatomy is deformed.
V DSC: We compared the Dice Similarity Coefficient (DSC) between the pre-liver mask and the deformed post-liver mask. The liver boundaries should match after DIR, so the DSC should be close to 1.0.
VI MAM: We compared the absolute differences in MAM measurements to assess the consistency of the three alternative methods with Morfeus, which has been proven to provide MAM in several retrospective studies^7,25,26^ and a prospective randomized Phase II clinical trial^2^. MAM is defined as the minimal distance between the tumor and ablation zone boundaries after registration. ≥ 5 mm is considered clinically adequate ^27^. In this study, we reported the absolute difference in MAM relative to Morfeus.
VII Efficiency: We compared computational efficiency using the average inference time per case.

We used the Wilcoxon signed-rank test with a threshold of 0.05 to assess the significance of performance differences between Morfeus, BioD, VFA, and VoxelMorph.

**Figure 3:**
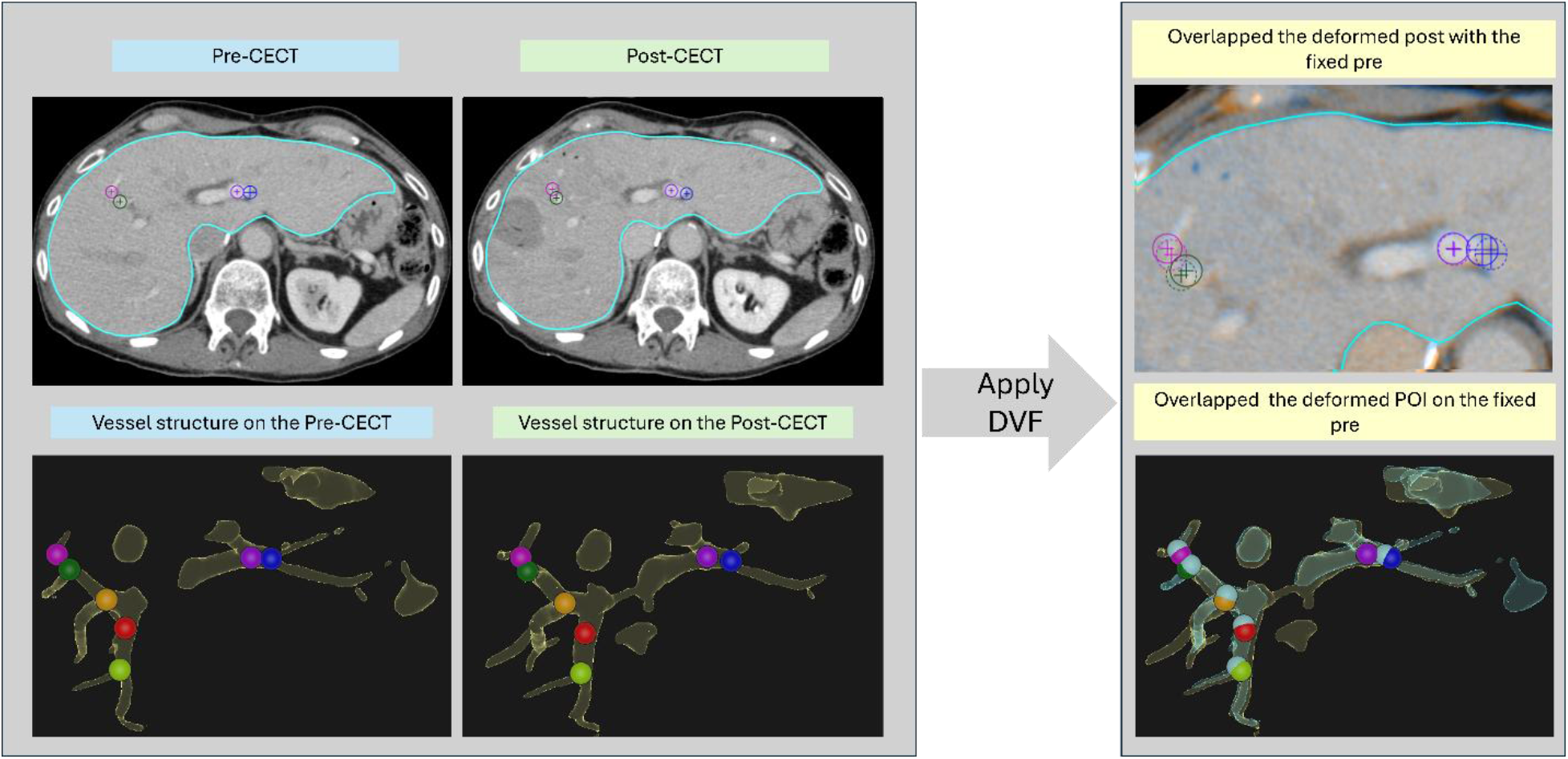
Example of bifurcation landmarks visualized on the pre-, post-CECT, and the deformed pre-CECT, registered using Morfeus-generated DVF and overlaid with the post-CECT. Top row: Solid-line circles are landmark positions on the post-CECT, and dashed-line circles are the corresponding deformed landmarks from the pre-CECT. Bottom row: Vessel structures with overlaid landmarks for each image. Seven distinct colors are used to represent the post-CECT landmarks, deformed pre-vessel structure shown as light blue, and the deformed pre-landmarks are shown as light blue spheres with a 1 cm diameter for visual clarity.

## Results

Our models were successfully trained and evaluated using the patient cohort summarized in Table 1.

**Table 1.** Patients’ demographic information.

| Dataset | Total |  |  | Testing |  |  |
| --- | --- | --- | --- | --- | --- | --- |
| Sex | Male | Femal | Total | Male | Female | Total |
| Count | 98 | 59 | 157 | 27 | 15 | 42 |
| Age (Mean, Min-Max) | 59.9 (21-83) | 54.5 (27-77) | 57.9 (21-83) | 63.1 (40-81) | 55.7 (27-81) | 60.5 (27-81) |
| Tumor Volume (Mean ± SD cc) | 2.5 ± 3.5 | 2.5 ± 2.7 | 2.5 ± 3.2 | 2.2 ± 2.5 | 1.5 ± 1.6 | 1.9 ± 2.2 |
| Number of colorectal liver metastases cases | 58 | 36 | 94 | 10 | 7 | 17 |
| Number of lesions (Median and Range) | 1 (1-5) | 1 (1-4) | 1 (1-5) | 1 (1-5) | 1 (1-3) | 1 (1-5) |
| Tumor in Segment 1 | 3 | 0 | 3 | 0 | 0 | 0 |
| Tumor in Segment 2 | 9 | 3 | 12 | 3 | 0 | 3 |
| Tumor in Segment 3 | 6 | 6 | 12 | 2 | 1 | 3 |
| Tumor in Segment 4 | 16 | 11 | 27 | 4 | 2 | 6 |
| Tumor in Segment 5 | 18 | 4 | 22 | 6 | 1 | 7 |
| Tumor in Segment 6 | 15 | 8 | 23 | 1 | 2 | 3 |
| Tumor in Segment 7 | 12 | 15 | 27 | 3 | 4 | 7 |
| Tumor in Segment 8 | 19 | 12 | 31 | 8 | 5 | 13 |

Note.— CRLM: colorectal liver metastases.

### Validation of BioDeformUNet Against the Morfeus Reference

We evaluate our BioDeformUNet method to assess whether its performance aligns with that of its guidance method, Morfeus. In Table 2, the tumor location test shows an average error of 1.84 mm, which is less than the one voxel diameter (3.0 mm). The displacement error exceeded 2mm in 8 cases (19%), 3mm in 8 cases (19%), and none of the errors exceeded 4 mm. In addition to local tumor performance, we also tested whole-liver performance, and it showed that, on average, 91.9% of the voxels have an error less than 3.0 mm, indicating that most of the registration differences between the BioDeformUNet and Morfeus are smaller than the size of an image voxel.

**Table 2:** Reference-based validation of BioDeformUNet relative to Morfeus using GTV centroid and whole-liver displacement metrics.

| Metric | BioDeformUNet vs Morfeus |
| --- | --- |
| Tumor-based metrics |  |
| Mean centroid distance (mm) | 1.84 ± 0.87 |
| Number of cases with centroid distance > 2 mm | 8 (19%) |
| Number of cases with centroid distance > 3 mm | 8 (19%) |
| Number of cases with centroid distance > 4 mm | 0 (0%) |
| Number of cases with centroid distance > 5 mm | 0 (0%) |
| Whole-liver-based metrics |  |
| Mean % voxels with displacement error < 3 mm | 91.90 ± 17.60 |

Note.—The table reports the average and standard deviation of the Euclidean distance between GTV
centroids obtained using BioDeformUNet and Morfeus (N = 43), the number of cases when increasing the error threshold, and the average percentage of whole-liver voxels with displacement magnitude error < 3mm (one voxel diameter).

The TRE test shows the average landmark errors of the BioDeformUNet and Morfeus (N=35). In Table 3, BioDeformUNet shows a TRE of 3.31 ± 1.17 mm, and Morfeus shows a TRE of 3.23 ± 1.06 mm. The Wilcoxon Rank-Sum test shows no significant difference between the two methods’ TRE test results (p-value = 0.41), suggesting close agreement.

**Table 3.** Tumor centroid error, TRE and DSC evaluation across VoxelMorph, VFA, BioDeformedUNet, and Morfeus.

| Method | Average tumor localization error (mm, N=43) | Average tumor size change (%<br>N=43) | Average TRE (mm, N=35) | Average DSC (%<br>N=43) |
| --- | --- | --- | --- | --- |
| VoxelMorph | 3.04 ± 1.73 | 20.00 ± 19.27 | 3.61 ± 1.28 | **98.85 ± 0.14** |
| VFA | 2.95 ± 1.26 | 12.00 ± 16.78 | **2.78 ± 0.96** | 93.83 ± 1.3 |
| BioDeformUNet | **1.84 ± 0.87** | **4.80 ± 3.90** | 3.31 ± 1.17 | 96.00 ± 1.3 |
| Morfeus | — | 4.75 ± 5.56 | 3.23 ± 1.06 | 96.58 ± 0.60 |

Note. — Tumor centroid error was calculated using the Euclidean distance between the Morfeus reference tumor location and the predicted tumor location. TRE was calculated using automatically identified bifurcations. Only cases with at least 3 landmarks were included (N=35). DSC was calculated using the pre-liver mask overlap with the deformed post-liver mask (N=43). The results are reported as mean ± standard deviation.

### Comparative Registration Performance Across Methods

We evaluated the tumor localization accuracy for VoxelMorph and VFA relative to Morfeus, and the results were shown in Table 3. The tumor centroid error of VoxelMorph relative to Morfeus was 3.04 ± 1.73 mm, which was significantly larger than BioDeformUNet (p-value < 0.001). Similarly, VFA showed a tumor centroid error of 2.95 ± 1.26 mm, which was also significantly larger than BioDeformUNet (p-value < 0.001) (Fig 4.A).

**Figure 4.**
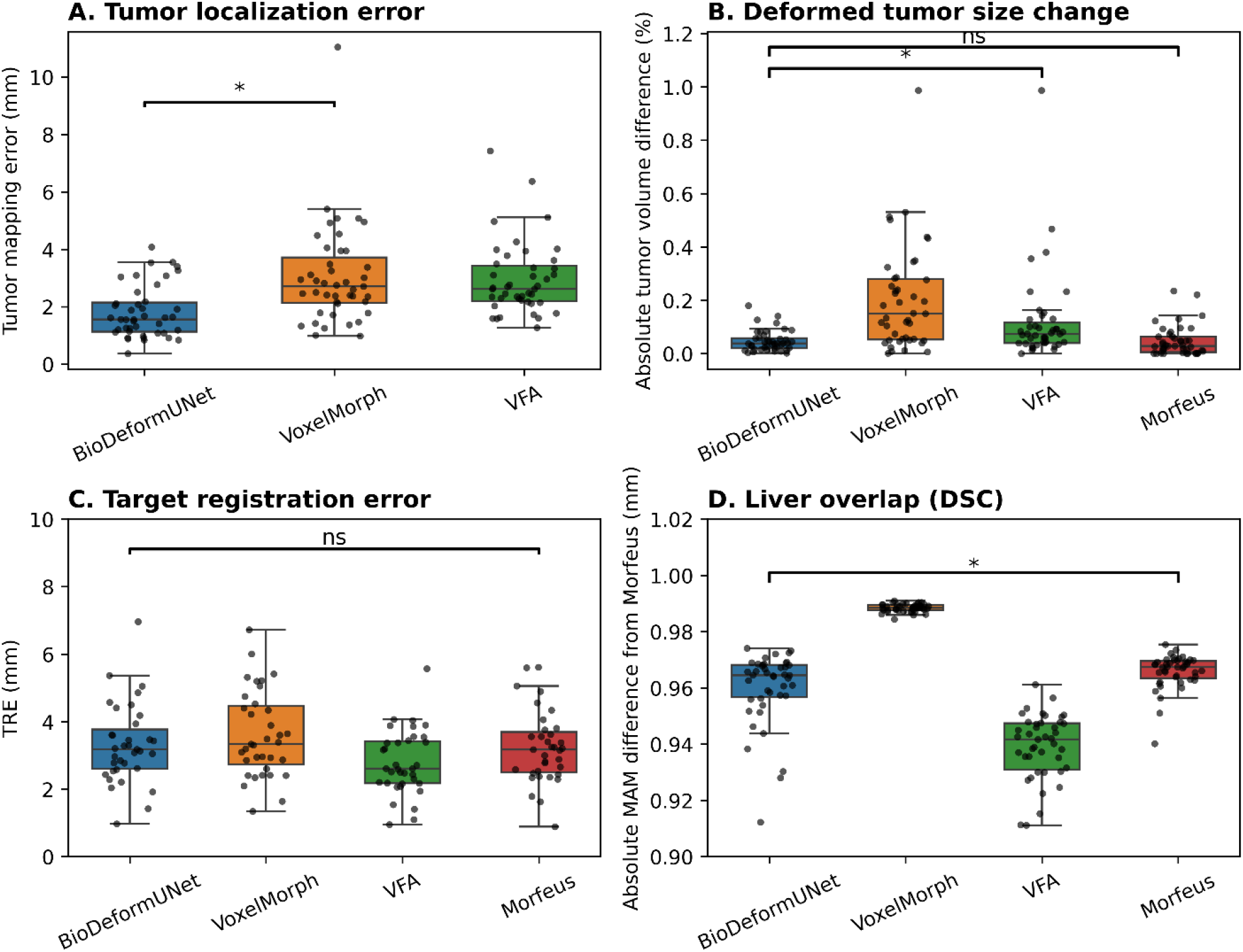
Evaluation results of different DIR methods. (A) Tumor localization error, (B) tumor volume change, (C) target registration error (TRE), and (D) liver overlap measured by Dice similarity coefficient (DSC) for BioDeformUnet (BioD), VoxelMorph, VFA, and Morfeus. Box plots show the median and IQR, and each dot represents an individual patient. BioD achieved results comparable to Morfeus in tumor localization accuracy, tumor volume preservation, and TRE, while demonstrating competitive performance relative to other deep learning methods. Liver DSC is included to evaluate the preservation of overall liver anatomy after deformation.

In addition to tumor localization accuracy, Table 3 shows changes in tumor volume before and after deformation. Morfeus showed a percentage tumor volume change of 4.75 ± 5.56%. BioDeformUNet showed a comparable change of 4.80 ± 4.01%, with no significant difference from Morfeus (p-value = 0.57). In contrast, VFA resulted in a significantly larger volume change than BioDeformUNet (12.00 ± 16.78%, p-value < 0.001). Similarly, for VoxelMorph, the tumor size change is significantly larger than BioDeformUNet (19.27 ± 20.00%, p-value < 0.001) (Fig 4.B).

The results of TRE tests for the VoxelMorph and VFA are shown in Table 3. It shows the average TRE of VoxelMorph was 3.61 ± 1.28 mm, which is significantly larger than BioDeformUNet (p-value = 0.005) and Morfeus (p-value < 0.001). VFA had a lower average TRE of 2.79 ± 0.96 mm, which is significantly smaller than BioDeformUNet (p-value < 0.001) and Morfeus (p-value < 0.001) (Fig 4.C).

Whole-liver registration performance was further evaluated by DSC, with the average DSC and standard deviation are shown in Table 3. VoxelMorph achieved significantly higher DSC than Morfeus (p-value < 0.001) and BioDeformUNet (p-value < 0.001). In contrast, VFA showed significantly lower DSC than Morfeus (p-value < 0.001) and BioDeformUNet (p-value < 0.001). BioDeformUNet also showed lower DSC than Morfeus (p-value < 0.001) (Fig 4.D).

The evaluation results of MAM consistency with Morfeus are summarized in Table 4. The absolute difference in MAM between BioDeformUNet and Morfeus is 0.4 [IQR: 0.1 – 0.7], which is significantly smaller than the difference between Morfeus and VoxelMorph (p-value = 0.009) and the difference between Morfeus and VFA (p-value = 0.002). In other words, the results indicate that BioDeformUNet shows a smaller absolute MAM difference from Morfeus than the other methods.

**Table 4.** MAM consistency related to Morfeus.

| Method | Absolute difference of MAM (mm) |
| --- | --- |
| VoxelMorph | 0.7 [0.1 - 1.3] * |
| VFA | 0.7 [0.1 - 1.4] * |
| BioDeformUNet | 0.4 [0.1 - 0.7] |

Note. —The MAM consistency was evaluated based on the absolute difference between Morfeus and the other three methods. The results were presented as median [interquartile range], and the two-sided Wilcoxon signed-rank test was used to assess the scientific significance compared with BioDeformUNet.
\* $p < 0.05$

The median runtime of BioDeformUNet to predict the DVF with one image pair is 0.59s (IQR: 0.57-0.60s). VoxelMorph required an average of 0.20s (IQR: 0.20-0.21s). VFA required an average of 0.33s (IQR: 0.33-0.34s). The historical observation runtime of Morfeus for the FEM and FEA component that is replaced with the proposed model is 20.20s (IQR: 17.40-23.25s). Overall, BioDeformUNet achieves comparable accuracy to the biomechanics-based DIR algorithm and is more time-efficient.

## Discussion

In this study, we developed a fully supervised 3D U-Net model, BioDeformUNet, for predicting DVFs from pairs of CT images in the context of liver ablation treatment. Our findings demonstrated that BioDeformUNet accurately predicts clinically relevant liver deformation and deformation within the tumor region, indistinguishable from the biomechanical reference model. BioDeformUNet exhibits efficient processing time and comparable accuracy, with balanced performance across different evaluation metrics.

BioDeformUNet achieved competitive TRE and DSC through supervised learning guided by biomechanical deformation targets. This guidance encourages anatomically consistent deformation, which enables local anatomical accuracy while preserving overall liver shape. Compared to the MSE loss function, which treats all voxels equally, the soft-weight ROI mask introduces spatially varying importance. Specifically, it gives greater attention to the liver region and to areas undergoing larger deformation. This design prioritizes clinically relevant areas and supports consistent deformation prediction in regions important for downstream MAM assessment.

We further compared the BioDeformUNet with two deep learning-based registration methods, VoxelMorph and VFA. VoxelMorph achieved the highest DSC, reflecting its ability to align global shapes, but also shows the highest TRE. VFA is an attention-based matching model that effectively captures local correspondence^15^ and demonstrates strong performance in deforming the vascular structure, resulting in the lowest TRE and lowest DSC across all evaluated methods. In contrast, BioDeformUNet shows more balanced performance across all evaluation metrics and achieves results comparable to those of the reference method, Morfeus. Accurate landmark deformation is important for the registration task; however, preserving global liver shape and maintaining consistent deformation across different regions are also important. Maintaining tumor volume is also important, as an unrealistic change in tumor volume can cause errors in downstream tasks such as MAM estimation. BioDeformUNet shows more stable volume preservation compared with VoxelMorph and VFA. Clinically, tumors can occur at different locations within the liver, including subcapsular, perivascular, and central regions. In our testing set, 24 tumors are in the subcapsular region. In such cases, having both accurate global alignment and stable local deformation is essential for reliable MAM estimation.

Accurate DVF estimation is important for reliable MAM assessment, as MAM assessments depend on tumor mapping from pre-CECT to post-CECT and measuring the distance to the ablation zone boundary. Small errors in the predicted deformation at the tumor boundary may alter the minimal distance and lead to errors in MAM assessment. In this study, BioDeformUNet was trained only to predict deformation between pre- and post-CECT images, and does not directly predict MAM, which requires additional geometric computation steps. However, BioDeformUNet shows a comparable accuracy as the reference method Morfeus in deformation estimation, supporting reliable downstream MAM assessment.

Beyond model accuracy, inference runtime is also important for clinical deployment. Morfeus typically takes about 20 seconds to process a single image pair. However, BioDeformUNet directly predicts DVF from the trained model, with an inference time of about 0.6 seconds, resulting in a substantial runtime reduction. More importantly, fundamentally BioDeformUNet does not require mesh generation from the segmentations, which is a further rate limiting steps in the Morfeus process requiring 60-180 seconds. Despite this significant speed advantage, BioDeformUNet maintains performance comparable to Morfeus. This improves efficiency and supports repeated assessments.

While BioDeformUNet demonstrated competitive performance, several limitations remain. First, BioDeformUNet is trained and evaluated using a validated biomechanics-based image registration method, Morfeus, and the computed DVF is used as a reference rather than a physical ground truth for deformation and MAM estimation. Obtaining physical ground-truth deformation is clinically difficult. Therefore, the reported results reflect relative agreement rather than absolute accuracy. Second, the relationship between computed MAM and clinical performance is not evaluated in this study, because it requires further investigation with follow-up data. Third, additional evaluation with challenging deformations is needed to test the model’s ability to handle extreme deformation.

This study explores deep learning’s ability to learn liver motion using ground-truth DVF data. The deep learning model can accurately and quickly predict deformation, providing a potential intraprocedural decision-making tool during percutaneous thermal ablation of liver cancers. In addition, the deep learning approach may enable greater algorithmic flexibility and the potential to overcome other constraints, such as the requirement to include the entire liver in the image. In the future, we intend to refine our approach by reducing data dimensionality, including full 3D image and limited-view 3D image registration, and full 3D image and 2D image registration, so that the image registration procedure during liver ablation can be more time-efficient.

## Acknowledgments

The National Cancer Institute of the National Institutes of Health supported the research under awards R01 CA235564, P30 CA016672. Dr. Noreen Siddiqi was supported by an Image Guided Cancer Therapy (IGCT) T32 Training Program Fellowship (T32CA261856). The research reported in this publication was supported in part by the resources of the Image Guided Cancer Therapy Research Program at The University of Texas MD Anderson Cancer Center through a generous gift from the Apache Corporation. We gratefully acknowledge the manuscript editing services provided by the Research Medical Library at The University of Texas MD Anderson Cancer Center. Copilot (Microsoft 365 based on the GPT-5 chat mode, Microsoft Corporation; accessed May 2025 to August 2026) and Grammarly AI (version 1.2.282.1933, Grammarly Inc.; accessed May 2025 to August 2026) were used only for grammatical correction and readability improvement. No AI was used for content generation, data analysis, or interpretation.

Dr. Kristy K. Brock reports grants from the US National Institutes of Health and Raysearch Laboratories; a licensing agreement with RaySearch Laboratories; travel support from the American Association of Physicists in Medicine and Raysearch Laboratories; and service on the clinical advisory board of Raysearch Laboratories. Dr. Bruno C. Odisio reports research grants from the National Institutes of Health, Siemens Healthineers, and Johnson & Johnson.

## Data sharing statement

Data in this study is not publicly available, but researchers may request access by contacting the authors.

