## Supplemental materials for "BioDeformUNet: A Deep Learning Model for Biomechanically Informed Liver Image Registration"

### Supplements

#### Supplement Section 1 - Rigid Registration in Data Preprocessing

The model may encounter new data that exhibit a large deformation due to the absence of rigid registration, and the currently available amount of data is insufficient to predict such a large deformation. A preliminary rigid registration step is applied to the data to enhance the model's performance before inputting it into the model. This rigid registration only involves translation adjustments.

The preliminary rigid registration was applied to the post-CECT and post-liver mask, ensuring that the liver's centroid in the post-CECT image aligns with its position in the pre-CECT image. The new DVF is generated by subtracting the translation displacement from the original DVF.

#### Supplement Section 2 – Loss Function Selection

We used 5-fold cross-validation to evaluate the 3 loss formulations: pure MSE, region-of-interest (ROI)-masked MSE, and magnitude-weighted ROI-masked MSE (the one reported in the main manuscript). Hyperparameters were selected exclusively using the validation folds, and the hold-out testing set was used for the final evaluation. For each formulation, patient-level validation results across all five folds are combined and shown in Table S1.

Table S1. Comparison of loss function

| Metric | Pure MSE | ROI masked | Magnitude-weighted | p-value: Pure MSE vs ROI masked MSE | p-value: Pure MSE vs Magnitude-weighted ROI-masked MSE | p-value: ROI masked vs Magnitude-weighted ROI-masked MSE |
|---|---|---|---|---|---|---|
| Tumor localization error (N=127, mm) | 2.29 ± 1.65 | **1.79 ± 1.53** | 1.94 ± 1.51 | <0.01 | <0.01 | 0.15 |
| Tumor size change (N=127, %) | 5.52 ± 5.62 | 5.00 ± 5.93 | **4.64 ± 5.18** | 0.37 | 0.11 | 0.55 |

| | | | | | | |
|---|---|---|---|---|---|---|
| TRE (N=55, mm) | 3.86 ± 1.64 | 3.66 ± 1.56 | **3.63 ± 1.54** | <0.01 | <0.01 | 0.32 |
| DSC (N=127, %) | 96.04 ± 0.94 | 95.91 ± 1.43 | 96.03 ± 1.33 | 0.54 | 0.02 | <0.01 |

Using a ROI-masked MSE loss function significantly improved GTV and TRE compared with using pure MSE. No statistically significant differences were observed between the ROI-masked MSE and magnitude-weighted ROI masked MSE losses for the tumor location test and TRE test. The magnitude-weighted ROI-masked MSE loss achieved a statistically significant improvement in DSC while maintaining comparable GTB and TRE performance and was therefore selected as the final model.

**Supplement Section 3 - Evaluation Matrices Details**

Tumor localization test:

The tumor centroid was defined as the center of mass of the tumor segmentation. We compared the geometric center of the deformed tumor, denoted $(x_{c_{true}}, y_{c_{true}}, z_{c_{true}})$ for the $DeformedTumor_{true}$ and $(x_{c_{pred}}, y_{c_{pred}}, z_{c_{pred}})$ for the $DeformedTumor_{pred}$, and then calculated the Euclidean distance between the centroids $ED_{T_c}$ of the ground truth and the predicted deformed tumors:

$$ED_{T_c} = \sqrt{\left(x_{c_{true}} - x_{c_{pred}}\right)^2 + \left(y_{c_{true}} - y_{c_{pred}}\right)^2 + \left(z_{c_{true}} - z_{c_{pred}}\right)^2}$$
